# Comparative performance of SARS CoV-2 lateral flow antigen tests demonstrates their utility for high sensitivity detection of infectious virus in clinical specimens

**DOI:** 10.1101/2021.02.27.21252427

**Authors:** Suzanne Pickering, Rahul Batra, Luke B. Snell, Blair Merrick, Gaia Nebbia, Sam Douthwaite, Amita Patel, Mark Tan Kia Ik, Bindi Patel, Themoula Charalampous, Adela Alcolea-Medina, Maria Jose Lista, Penelope R. Cliff, Emma Cunningham, Jane Mullen, Katie J. Doores, Jonathan D. Edgeworth, Michael H. Malim, Stuart J.D. Neil, Rui Pedro Galão

## Abstract

**Background:** Rapid antigen lateral flow devices (LFDs) are set to become a cornerstone of SARS-CoV-2 mass community testing. However, their reduced sensitivity compared to PCR has raised questions of how well they identify infectious cases. Understanding their capabilities and limitations is therefore essential for successful implementation. To address this, we evaluated six commercial LFDs on the same collection of clinical samples and assessed their correlation with infectious virus culture and cycle threshold (Ct) values.

**Methods:** A head-to-head comparison of specificities and sensitivities was performed on six commercial rapid antigen tests using combined nasal/oropharyngeal swabs, and their limits of detection determined using viral plaque forming units (PFU). Three of the LFDs were selected for a further study, correlating antigen test result with RT-PCR Ct values and positive viral culture in Vero-E6 cells. This included sequential swabs and matched serum samples obtained from four infected individuals with varying disease severities. Detection of antibodies was performed using an IgG/IgM Rapid Test Cassette, and neutralising antibodies by infectious virus assay. Finally, the sensitivities of selected rapid antigen LFTs were assessed in swabs with confirmed B.1.1.7 variant, currently the dominant genotype in the UK.

**Findings:** Most of the rapid antigen LFDs showed a high specificity (>98%), and accurately detected 50 PFU/test (equivalent N1 Ct of 23.7 or RNA copy number of 3×10^6^/ml). Sensitivities of the LFDs performed on clinical samples ranged from 65 to 89%. These sensitivities increased in most tests to over 90% for samples with Cts lower than 25. Positive virus culture was achieved for 57 out of 141 samples, with 80% of the positive cultures from swabs with Cts lower than 23. Importantly, sensitivity of the LFDs increased to over 95% when compared with the detection of infectious virus alone, irrespective of Ct. Longitudinal studies of PCR-positive samples showed that most of the tests identified all infectious samples as positive, but differences in test sensitivities can lead to missed cases in the absence of repeated testing. Finally, test performance was not impacted when re-assessed against swabs positive for the dominant UK variant B.1.1.7.

**Interpretation:** In this comprehensive comparison of antigen LFD and virus infectivity, we demonstrate a clear relationship between Ct values, quantitative culture of infectious virus and antigen LFD positivity in clinical samples. Our data support regular testing of target groups using LFDs to supplement the current PCR testing capacity, to rapidly identify infected individuals in situations where they would otherwise go undetected.

**Funding:** King’s Together Rapid COVID-19, Medical Research Council, Wellcome Trust, Huo Family Foundation.

## Introduction

Covid-19 continues to have a profound impact on global health, with many countries resorting to economically and socially damaging restrictions to minimise the spread of SARS-CoV-2 and protect healthcare systems from being overwhelmed.

Pathways out of national lockdowns - and strategies to mitigate the need for them in the future – depend upon the successful implementation of mass vaccination programmes, effective contact tracing systems and mass community testing. In addition to the existing PCR-based testing systems, the latter may take the form of targeted intensive testing in increasing incidence areas, alongside regular routine screening in healthcare, education, workplace and leisure settings. Realistically, the expansion of mass regular testing relies heavily on an element of low-infrastructure- or self-testing, such as that offered by rapid antigen lateral flow devices (LFDs).^1,2^

Thoroughly understanding the advantages and limitations of rapid antigen LFDs is, therefore, a priority and will help to inform decisions about where these tests will have the most utility and, conversely, where they could be contraindicated. There are concerns about their reduced sensitivity in comparison to PCR, and controversies have arisen over the suitability of their implementation.^3–5^ Problems with comparing Ct values from RT-qPCR between different protocols, and even between the same protocols at different locations, combined with uncertainty about the range of viral loads that constitute a transmission risk, have been the root of many of the issues.^5,6^ Individuals are most infectious around the time of symptom onset, when viral loads in the upper respiratory tract are highest,^7,8^ with recent studies confirming an association between viral load and increased transmission of SARS-CoV-2.^9^ For asymptomatic individuals, infectivity and viral load dynamics involve a similar, limited, period of infectivity, and asymptomatic and pre-symptomatic contributions to spread in the community remain problematic.^10,11^

Several studies have shown a relationship between Ct value and virus infectivity,^8, 12–14^ and manufacturers of rapid antigen LFDs have implied a link between antigen test positivity and infectious potential. Here we present a detailed assessment of the relationship between Ct, quantitative culture of infectious virus and antigen test positivity, alongside an independent and unbiased head-to-head comparison of six widely available commercial antigen tests. Most tests showed good sensitivity (>90%) at Cts of less than 25, with sensitivity increasing to over 95% when compared to infectious samples. Longitudinal studies of PCR-positive samples highlight the importance of regular testing. We also re-assessed the tests against the dominant genotype in the UK, B.1.1.7, and found no difference in test performance.

## Materials and Methods

### Study samples and ethics

Combined nasal/oropharyngeal swabs were submitted to the diagnostic laboratory in 1 ml of viral transport medium (VTM) for routine real time SARS-CoV-2 RT-PCR testing. Surplus VTM was routinely stored at −80°C by the diagnostic laboratory for future technology evaluations. VTM from 100 laboratory-confirmed SARS-CoV-2 positive swabs selected to cover a wide range of Cts (14 to 39) and 100 confirmed negative swabs were used for head-to-head comparisons of 6 commercial antigen tests. Similarly, VTM from an additional 141 confirmed positive swabs were used for comparative studies on infectivity and antigen testing. All samples were collected between March and October 2020. A further 23 laboratory-confirmed SARS-CoV-2 positive swabs, collected in January 2021, were shown by on-site whole-genome sequencing to be from the B.1.1.7 variant and used for comparative evaluation of the sensitivity of selected tests.

Matched routinely-collected serum samples stored for up to 48 hours at 4°C in the Viapath Blood Sciences laboratory were retrieved after routine diagnostic testing prior to planned discard and stored at −80°C for future serological analysis. All swabs, VTM and serum samples were stored in the Directorate of Infection. Samples for research were retrieved by the primary care team and anonymised before sending to the King’s College London laboratories for analysis along with dates of symptom onset and sample collection, and any relevant routine laboratory result obtained from that sample. All studies were performed in accordance with the UK Policy Framework for Health and Social Care Research and with specific Research Ethics Committee approval (REC 20/SC/0310).

### Viral growth assays

For the comparative studies on infectivity and antigen positivity, each swab was subjected to the following procedures: RNA extraction for subsequent RT-PCR and sequencing; titration and viral titre determination by plaque assay; titration and infectivity determination by intracellular anti-N staining; viral propagation for isolation of virus; and rapid antigen testing. Viral growth assays were performed on Vero.E6 cells. For plaque assays, VTM was 10-fold serially diluted and applied to Vero.E6 cells in 12-well plates, in a volume of 500 ul per well, and incubated for 1 hour at 37°C. 500 ul of pre-warmed overlay (0.1% agarose in DMEM supplemented with 2% FCS, pen/strep and amphotericin B) was then applied to each well, and cultures were incubated for 72 hours at 37°C, before fixing with 4% formaldehyde. A solution of 0.05% crystal violet in ethanol was applied to each well, incubated for 5 minutes at room temperature, before washing with PBS, air drying and counting plaques.

### RT-PCR

Initial diagnostic laboratory testing was carried using the AusDiagnostics Multiplex Tandem SARS-CoV-2 PCR assays at Viapath Infection Sciences laboratory, St Thomas’ Hospital, London, and positive or negative swabs were selected on basis of this diagnostic test. For additional PCR testing and to ensure uniformity of RT-PCR conditions and Ct determination, RNA was extracted from 100ul of swab using the Qiagen QIAamp Viral RNA Kit following manufacturer’s instructions and eluted in 60ul of water. RT-PCR reactions (total volume 20µl) were performed with 5µl of eluted RNA, 4x TaqMan Fast Virus 1-Step Master mix (Applied Biosystems) and CDC’s IDT Primer-Probes Sets targeting SARS-CoV-2 N gene regions or human RNAse P, using a QuantStudio 5 (ThermoFisher Scientific). RNA standards were extracted as above from serial dilutions of a NATtrol™ SARS-CoV-2 Stock (ZeptoMetrix), which is constituted of inactivated intact viral particles with known RNA viral load.

### Rapid antigen tests

Tests were performed according to the manufacturers’ instructions, with the exception that swabs stored in viral transport medium (VTM) were used for evaluations, rather than direct swabs performed immediately prior to test performance. 50 ul of stored swab was mixed with 100 ul of buffer supplied with the test kit, and 100 ul of this was applied to the test cassette. Results were scored at the time stipulated by the manufacturer (between 10 and 30 minutes). Results were recorded independently by two readers, compared, and in the event of a discordant score were referred to a third individual. For the purpose of comparison, chromatographic tests were scored according to whether the test band was strongly positive (2), clearly positive (1), weakly positive (0.5) or negative (0).

### Neutralisation assays

Neutralisation assays were performed on Vero.E6 cells with replication-competent SARS-CoV-2 (England 02/2020) as previously described.^15^ Briefly, 20,000 cells were seeded per well of a 96-well plate the day before assay. Heat-inactivated sera were 3-fold serially diluted and incubated with 400 PFU per well of SARS-CoV-2 for 1 hour at 37°C. Medium was removed from the cells and replaced with the virus/serum mixtures. After 24 hours at 37°C, cells were fixed in 4% formaldehyde before intracellular nucleocapsid staining.

### Intracellular nucleocapsid staining

Immunostaining for SARS-CoV-2 nucleocapsid detection in Vero.E6 cells was performed *in situ* in formaldehyde-fixed 96-well plates, to verify viral culture experiments and as a read-out for infectious virus neutralisation assays, as described previously.^15^ Briefly, cells were permeabilised with 0.1% triton in PBS for 15 minutes, then blocked in 3% milk for 15 minutes at room temperature. Primary antibody (murinized anti-N 3009) was incubated at a final concentration of 2 ug/mL in 1% milk for 45 minutes at room temperature, before washing twice with PBS and incubating with secondary antibody (goat-anti-mouse IgG HRP-linked, Cell Signaling Technology, 1:2000) in 1% milk for 45 minutes at room temperature. Cells were washed twice with PBS, before addition of substrate. For SARS-CoV-2 plaque verification assays, TrueBlue HRP substrate was used (Seracare Life Sciences Inc.); for neutralisation assays, 1-Step Turbo TMB-ELISA substrate (ThermoFisher Scientific) was applied to the cells before quenching with sulfuric acid and reading at 450 nm.

### Rapid antibody tests

Presence of IgM and IgG antibodies in matched serums was assessed using the lateral flow immunoassay (LFIA) COVID-19 IgG/IgM rapid test cassette (SureScreen). 10 μl of serum was added to the LFIA membrane start point, followed by 2 drops of supplied buffer. Kits were run at room temperature for 10 minutes and scored both IgM and IgG. Scoring (negative, borderline, positive, strong positive) was performed independently by two individuals.

### Statistical Analyses

Expected binomial exact 95% confidence intervals were calculated on Prism 8.0 using Wilson/Brown statistical analysis. Significance of comparative sensitivities using swabs from different time periods was determined by repeated measures ANOVA.

## Results

### Head-to-head comparison of specificities, sensitivities and limits of detection of commercial rapid antigen tests

Six commercial rapid antigen tests (Innova, E25 Bio, SureScreen visual (V), Spring, Encode and SureScreen fluorescent (F); **Table 1**) were compared in specificity, limit-of-detection (LOD) and sensitivity validations.

**Table 1:** Rapid antigen LFD names and manufacturers.

| Rapid Antigen LFD | Distributor/Manufacturer | Reference |
| --- | --- | --- |
| SARS-CoV-2 Antigen Test Kit | Innova Med Group | N/A |
| SARS-CoV-2 Antigen Rapid Test Cassette (Swab) | Spring Healthcare | SP-SW 106 |
| SARS-CoV-2 Antigen Rapid Test Cassette (Nasopharyngeal Swab) | SureScreen Diagnostics Ltd | COVID19 AGVCT |
| SARS-CoV-2 Antigen Rapid Test Device | Emmo Pharma / Encode | N/A |
| COVID-19 Antigen Rapid Fluorescent Cassette | SureScreen Diagnostics Ltd | COVID19 AGC |
| Rapid Diagnostic Test | E25Bio | N/A |

Comparative specificity was determined for each test using a panel of 100 SARS-CoV-2 negative swabs (**Table 2**). All tests demonstrated high (>98%) specificity, with the exception of E25Bio. SureScreen and Encode achieved 100%. None of the negative samples gave false positive results in more than one test, suggesting that false positives appear stochastically and are not a particular feature of the samples. All false positives were only weakly positive, with the exception of SureScreen F for which this information was not available as the electronic reader delivers a binary result.

**Table 2:**
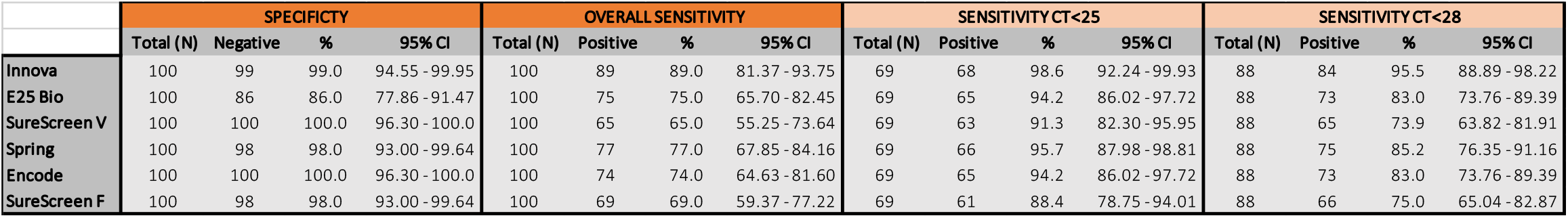
Comparative specificity and sensitivity of six commercial SARS-CoV-2 rapid antigen tests.

LODs were determined using specified plaque-forming units (PFU) of Vero.E6-propagated SARS-CoV-2 (England 02/2020 strain). Serial dilutions of virus stock were applied to each test in triplicate. Informed by the specificity determinations - in which there were very few false positives - any visible band was considered positive regardless of intensity or relationship to the control band. Although the tests are qualitative, results are displayed as a heatmap to convey the magnitude of the result, allowing more detailed comparisons between the tests and potentially informing future use. Most tests reliably detected 50 PFU per test (1500 PFU per mL) with the exception of Encode and SureScreen F (**Figure 1A**). SureScreen V and Innova had the lowest consistent LOD, and upon further testing SureScreen also consistently detected 20 PFU per test (600 PFU/mL; data not shown). Calibration experiments conducted on SARS-CoV-2 laboratory stocks (**Figure 1B**) and standardised RNA control reagents (**Figure 1C**) delivered the equivalent N1 Ct of 23.7 or RNA copy number of 3×10^6^/ml for the LOD of 1500 PFU per mL. As particle-to-infectious unit ratios can vary between viral variants or according to growth or assaying conditions, as an additional control we applied the Zeptometrix NATrol inactivated viral particle standard to the two tests with the best LOD – Innova and SureScreen. This was weakly positive on both tests at a copy number of 1.2×10^6^/ml (data not shown), or projected Ct value of 25, in agreement with the results shown in **Figure 1**.

**Figure 1.**
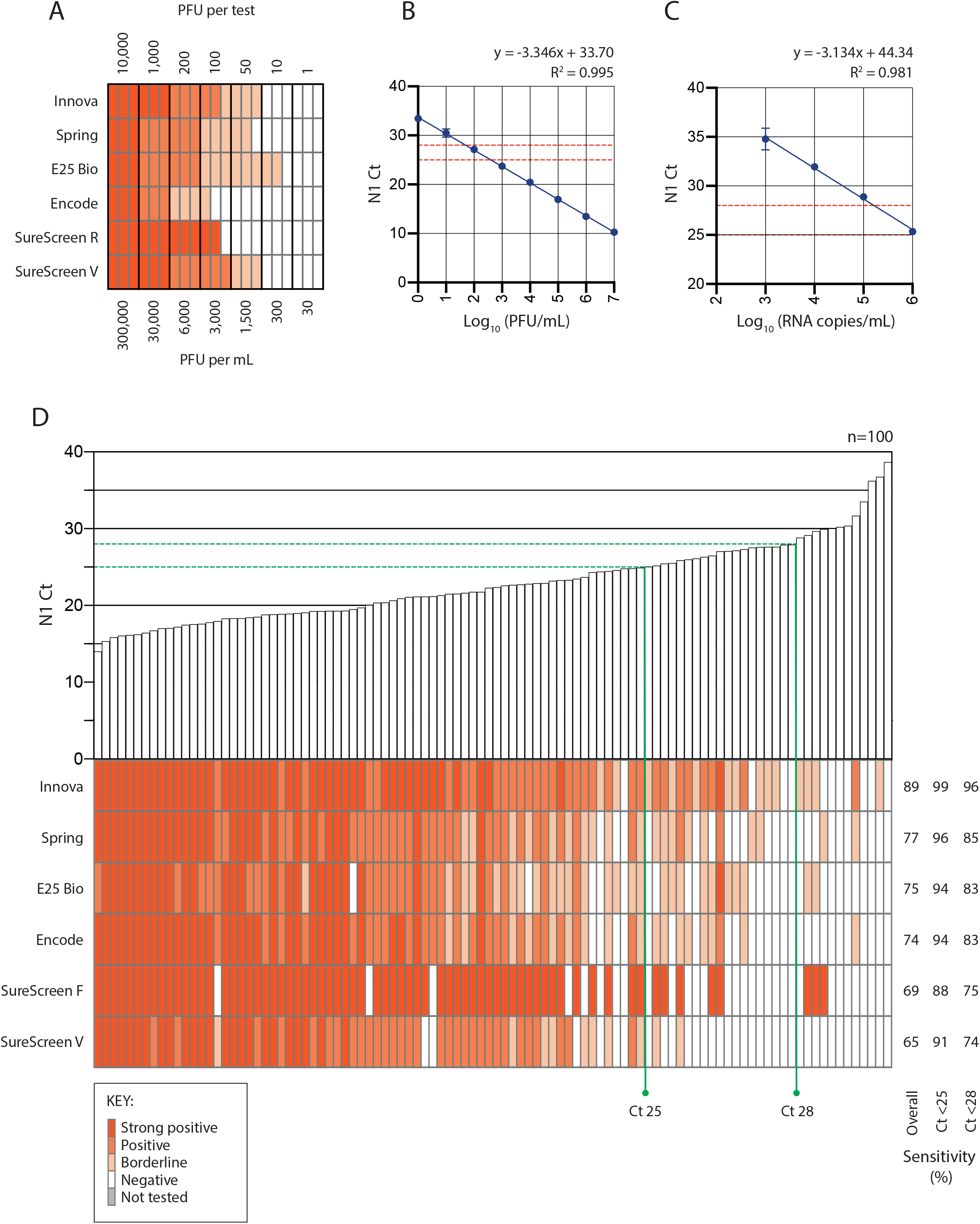
Comparative sensitivity of six commercial SARS-CoV-2 rapid antigen tests. (A) Limits of detection were compared for six commercial SARS-CoV-2 rapid antigen tests (Innova, Spring, E25 Bio, Encode, SureScreen visual (V) and SureScreen fluorescent (F)). 1-10,000 PFU of Vero.E6-propagated SARS-CoV-2 (England 02/2020) were applied to each test in triplicate (equating to 30-300,000 PFU/mL). For the purpose of comparison the results are scored according to whether the test band was strongly positive (score = 2), unequivocally positive (1), weakly positive (0.5) or negative (0), and presented as a heatmap. Equivalent Cts are given in (B). (B) Association between PFU/mL and threshold cycle (Ct) result for RT-PCR for the SARS-CoV-2 *n* gene (N1 Ct). RNA was extracted from serial dilutions of Vero.E6-titred SARS-CoV-2 (England 02/2020) and assayed by N1 RT-PCR. Error bars represent SD of three independent experiments. Points were fitted with a semi-log regression, shown on the graph, with the equation and the R^2^ value given above the graph. Horizontal dashed red lines denote Ct values of 25 and 28, the threshold cut-offs used for sensitivity determinations in (D). (C) Association between RNA copy number and N1 Ct values. Copy number/mL was derived from N1 RT-PCRs conducted on the Zeptometrix RNA standard. Points were fitted with a semi-log regression, shown on the graph, with the equation of the line and the R^2^ value given above the graph. Horizontal dashed red lines denote Ct values of 25 and 28, the threshold cut-offs used for sensitivity determinations in (D). (D) Tests were evaluated in head-to-head comparisons on an identical panel of 100 SARS-CoV-2-positive nasal/oropharyngeal swabs. Bars denote the N1 Ct result for each swab, shown in ascending order, and the antigen test results for each sample are shown directly below each bar. As for (A), antigen test results are presented as a heatmap. Sensitivity determinations from this sample set are shown to the right of the heatmap for each test. Thresholds of Ct 25 and 28, corresponding to 1.5×10^6^ and 1.65×10^5^ RNA copies/mL, or 400 and 50 PFU/mL, respectively, are indicated on the figure as vertical green lines with corresponding sensitivity values for each test at each threshold shown to the right of the heatmaps.

Sensitivity comparisons on clinical samples were performed as head-to-head evaluations on 100 PCR-confirmed SARS-CoV-2 combined nasal/oropharyngeal swabs with Cts ranging from 14 to 39 (**Figure 1D**). Swabs were selected to cover a wide range of Cts, from 14 to 39. Overall sensitivities are presented, as well as sensitivities determined for samples with a Ct less than 25, corresponding to approximately 1.5×10^6^ copies/ml or 400 PFU/ml, and Ct 28, corresponding to a copy number of approximately 1.65×10^5^ copies/ml or 50 PFU/ml. Innova had the highest overall sensitivity (89%) using clinical samples, with this increasing to 96 and 99% when applied to samples with Cts of less than 28 and 25 respectively (**Table 2**). All other tests had overall sensitivities of between 65 and 77%, increasing to over 90% for samples with Cts of less than 25 for all tests except SureScreen F. Thus, there was good sensitivity and specificity for all tests on swabs within a defined Ct window.

### Select LFDs predict swabs with infectious culturable SARS CoV-2 with very high sensitivity

Three of the rapid antigen tests from the first phase of comparisons were then selected for more detailed comparisons: the test with highest sensitivity (Innova), highest specificity (Encode) and a test with alternative technology (fluorescent machine-read result; SureScreen F). 141 combined nasal/oropharyngeal swabs were compared for N1 Ct value, antigen test result, and positive viral culture (**Figure 2A**). Samples covered a range of Cts from 12.7 to 40. The direct viral titre of the swabs was determined by plaque assay of serially diluted swabs, with additional confirmatory intracellular anti-SARS-CoV-2 nucleocapsid staining performed on viral culture samples for 110 of the 141 samples. 57 of the 141 samples were positive for viral growth (40.4%). 80% of cultures yielded Cts of less than 23, and the highest Ct from a sample with positive viral culture was 26.3. The latest time point that we were able to isolate virus from was 15 days post onset of symptoms. Titres of infectious virus in the samples showed an inverse linear relationship with N1 Ct values (**Figure 2B**). Both viral culture and antigen test positivity were associated with Ct value, rather than the timing of the sample post onset of symptoms (**Figure 2C**).

**Figure 2.**
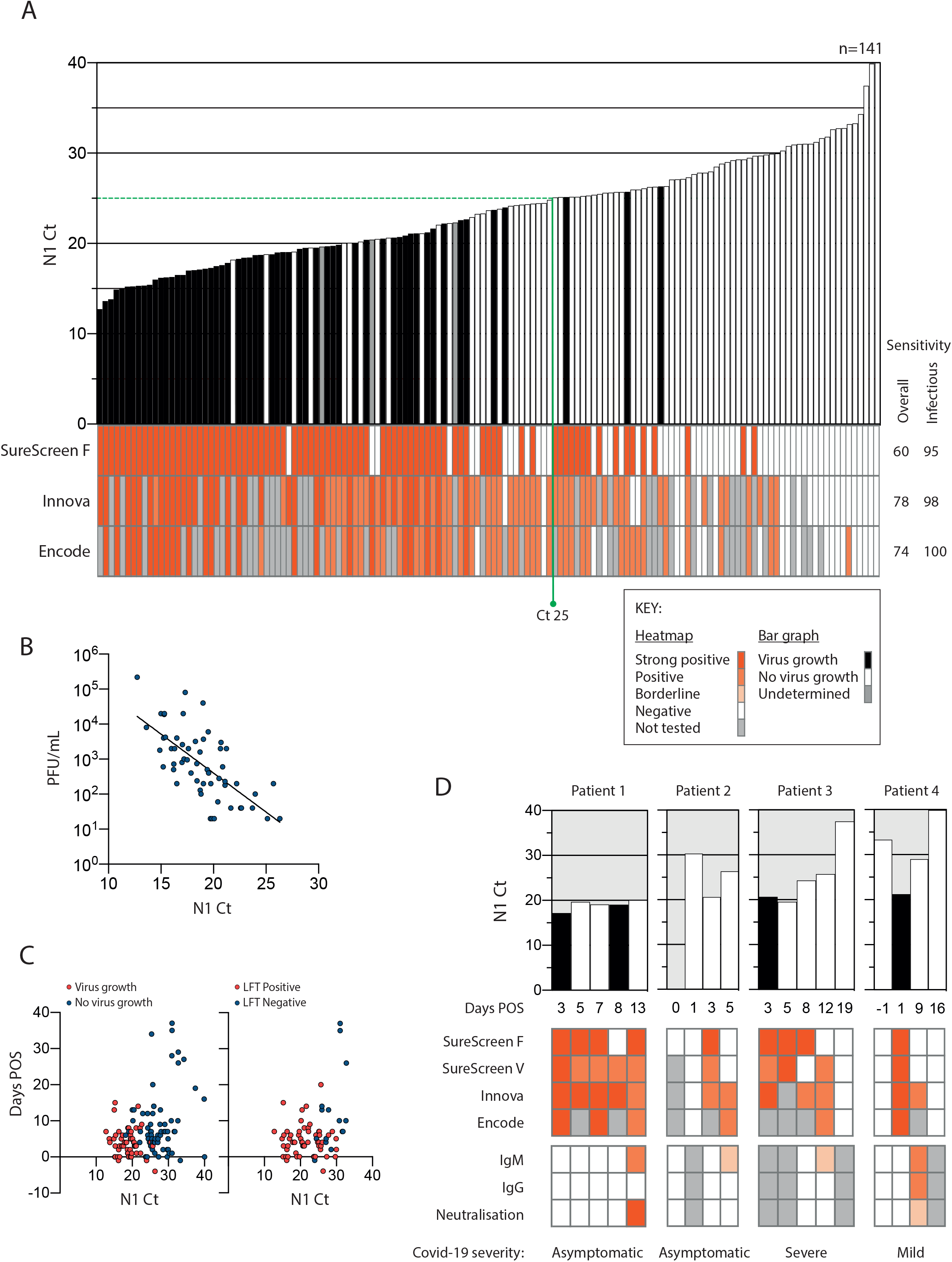
Comparison of Ct value, infectivity and rapid antigen test result for 141 clinical samples and four longitudinal examples. (A) Infectivity, SARS-CoV-2 N1 Ct, and antigen test results for 3 commercial tests were determined for 141 combined nasal/oropharyngeal swabs. Bars show the N1 Ct result for each swab, shown in ascending order, and are individually coloured according to whether virus was cultured from the sample. Black indicates successful virus isolation, white indicates no virus growth, and grey undetermined. Antigen test results for each sample are shown directly below each bar. As for Figure 1, results are scored according to whether the test band was strongly positive (score = 2), unequivocally positive (1), weakly positive (0.5) or negative (0), and presented as a heatmap; grey boxes indicate samples that were not tested on that particular test. Sensitivity determinations from this sample set are shown to the right of the heatmap for each test, showing overall sensitivity and sensitivity relative to infectious samples. (B) Direct viral titres of swabs was determined by plaque assay and compared with N1 Ct value. (C) Ct results from (A) plotted against days post onset of symptoms for each sample. Points are coloured according to whether virus growth was observed (red) or not (blue), left graph, or whether the Innova antigen test was positive (red) or negative (blue), right graph. (D) Longitudinal examples of infectivity, antigen test positivity, Ct and antibody detection. Sequential combined nasal/oropharyngeal swab samples and matched serum samples were obtained from four SARS-CoV-2-infected individuals. As for (A), bars show the N1 Ct value for each sample, shaded according to whether samples were virus culture positive (black) or negative (white). Antigen test results for each sample are shown below the bars. Antibody detection in matched serum samples are shown in the bottom panels, with heatmaps representing the magnitude of IgM and IgG detection in the SureScreen Covid-19 IgG/IgM Rapid Test Cassette or the detection of neutralising antibodies to SARS-CoV-2 in an infectious virus neutralisation assay on Vero.E6 cells.

Overall antigen test sensitivities and rankings were similar to those seen in **Figure 1**, with Innova delivering the highest sensitivity at 78%, then Encode (74%), followed by SureScreen F (60%) (**Table 3**). When compared only to the samples from which virus was cultured, all tests achieved a sensitivity of at least 95% (**Figure 2A** and **Table 4**). One point to note is that due to sample volume or test availability, not all samples could be assayed on Innova and Encode.

**Table 3:**
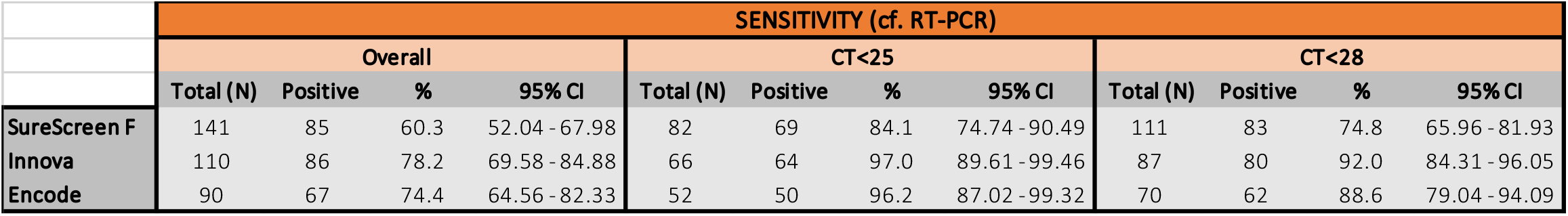
Sensitivity of three commercial SARS-CoV-2 rapid antigen tests compared with RT-PCR.

**Table 4:** Sensitivity of three commercial SARS-CoV-2 rapid antigen tests compared with Virus Isolation.

|  | SENSITIVITY (cf. Virus Isolation) |  |  |  |  |  |  |  |  |  |  |  |
| --- | --- | --- | --- | --- | --- | --- | --- | --- | --- | --- | --- | --- |
|  | Overall |  |  |  | CT<25 |  |  |  | CT<28 |  |  |  |
|  | Total (N) | Positive | % | 95% CI | Total (N) | Positive | % | 95% CI | Total (N) | Positive | % | 95% CI |
| SureScreen F | 57 | 54 | 94.7 | 85.63 - 98.57 | 54 | 52 | 96.3 | 87.46 - 99.34 | 57 | 54 | 94.7 | 85.63 - 98.57 |
| Innova | 46 | 45 | 97.8 | 88.66 - 99.89 | 43 | 42 | 97.7 | 87.94 - 99.88 | 46 | 45 | 97.8 | 88.66 - 99.89 |
| Encode | 34 | 34 | 100.0 | 89.85 - 100.0 | 32 | 32 | 100.0 | 89.28 - 100.0 | 34 | 34 | 100.0 | 89.85 - 100.0 |

To investigate the change of antigen test result over time, sequential combined nasal/oropharyngeal swabs were retrieved where available from 4 infected inpatients with varying disease severities and compared for N1 Ct value, antigen test result (SureScreen F, SureScreen V, Innova and Encode), and positive viral culture (**Figure 2D**). Matched serum samples were also retrieved from the same individuals where routine testing had been performed, and assayed for the appearance of antibodies (IgM and IgG by SureScreen COVID-19 IgG/IgM Rapid Test Cassette), and neutralising antibodies by infectious virus assay). All tests identified infectious samples as positive (with the exception of one sample by SureScreen F), and continued to deliver positive results for several days after peak infectivity. In patients 2-4, Innova (and where available, Encode) tested positive for up to 8 days longer than SureScreen F, although the exact length of this extended positivity cannot be stated as intermediate samples were not obtained. In two cases, PCR testing identified pre-infectious individuals (Patients 2 and 4) who were negative in all antigen tests. Two days later, a drop in Ct value coincided with antigen test positivity for all tests and the isolation of infectious virus in one of these individuals. Longitudinal results therefore highlight the importance of repeat, rather than one-off, testing with rapid antigen LFDs.

In two individuals, the occurrence of neutralising antibodies coincided with a dip in antigen test band strength (**Figure 2D**; patients 1 and 4). We also examined matched serum samples from 76 of 141 samples shown in Figure 2A and found no relationship between antigen test positivity, infectivity and the presence of antibodies (data not shown).

### Rapid antigen tests detect the B.1.1.7 variant with equivalent sensitivity

Given that the rapid antigen tests rely on antibody detection of SARS-CoV-2 nucleocapsid (N), even single amino acid mutations have the potential to impact test sensitivity. As such, test performance should be re-assessed in light of new emerging variants of SARS-CoV-2, such as B.1.1.7 that has become the dominant genotype in the UK and contains four mutations in N compared to England 02/2020 (D3L, R203K, G204R and S235F).^16^ We performed small-scale evaluations using matched swab samples spanning a range of Cts (12.7 to 31.8), from April 2020, September 2020, and confirmed (by viral sequencing) B.1.1.7-positive swabs from January 2021 (**Figure 3**). Both Innova and SureScreen V tests showed variations in sensitivities between the three batches of samples tested, as would be expected for biological samples, but there was no evidence of altered sensitivity for the B.1.1.7 swabs.

**Figure 3.**
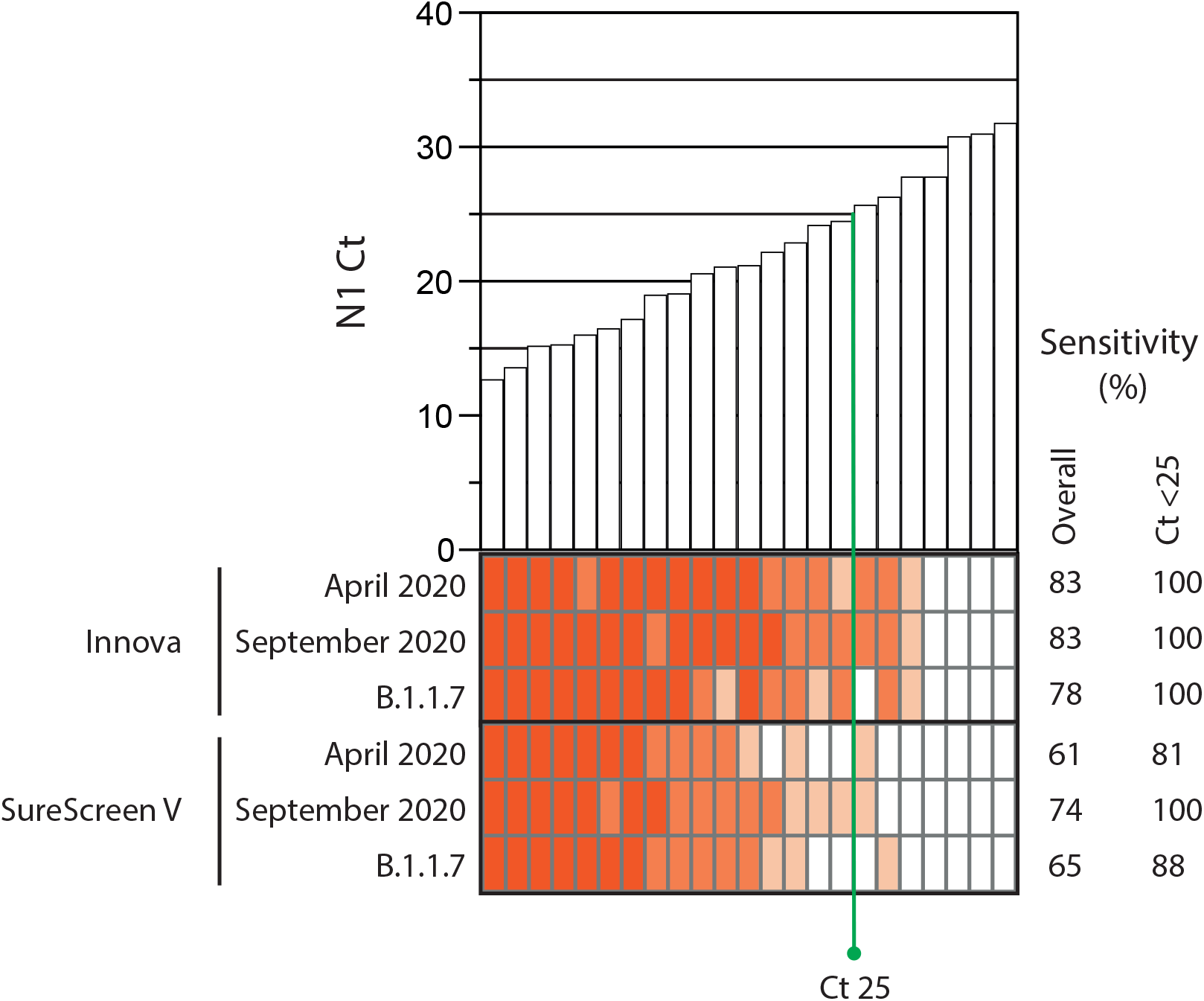
Comparative evaluation of antigen test sensitivity for the B.1.1.7 variant vs. England 02. Combined nasal/oropharyngeal swabs were obtained from 23 individuals with confirmed SARS-CoV-2/B.1.1.7 infection and compared with samples from before the variant was widely circulating in the UK population - May and September 2020. All swabs were matched for N1 Ct values, shown in ascending order in the bar graph, and tested on Innova and SureScreen visual rapid antigen tests. The Ct 25 threshold, corresponding to 1.5×10^6^ RNA copies/mL or 400 PFU/mL, is indicated in green.

## Discussion

Through extensive head-to-head comparison, we found that most rapid antigen tests performed to a high standard, with good sensitivity and excellent specificity. Consistent with previous reports, the tests delivered an overall sensitivity of 65 to 89% in comparison with PCR,^17–20^ rising to over 90% for most tests when compared with samples with Cts of 25 or less.^18, 21,22^ Sensitivity levels increased to over 95% when compared with samples that were infectious *in vitro*, with direct viral titre in the specimens correlating with Ct levels. This is the most comprehensive comparison of antigen LFD and infectivity to date, demonstrating a clear relationship between Ct values, quantitative culture of infectious virus and antigen LFD positivity, with all tests delivering reliable identification of infectious clinical samples.

In agreement with previous studies, we cultured virus from upper respiratory tract specimens with Cts of up to 26 (an equivalent viral load of 7×10^5^ copies/mL),^13,14,23^ with the majority of the culturable samples taken in the first week following symptom onset.^12–14, 24^ The minimum viral titre required for transmission is unclear^24^ and will depend in part on the proximity and duration of contact. Nevertheless, it has been reported that higher viral loads, as measured by lower Ct, are strongly associated with transmission^9^ and therefore reasonable to assume that the quantity of cultured virus *in vitro* has a similar comparable correlation with infectivity. There are, however, problems attempting to standardise Ct values as surrogate measures of potential to transmit, due to differences in RNA extraction and RT-PCR methods. This is demonstrated by a recent study in which differences of greater than 5 Cts between RT-PCR systems prompted fears that rapid antigen LFDs were missing up to 50% of potentially infectious cases.^4–6^ There have been frequent suggestions for SARS-CoV-2 results to be presented as viral load (copies/mL) due to difficulties in comparing Ct values between studies. However, the lack of an agreed standard for determining viral load is itself a problem, with reported viral loads often appearing even more disparate than Cts.^25^

While the reduced sensitivity of LFDs relative to PCR is less of a concern late in the infection course when Cts are rising and the risk of onward transmission is negligible,^9^ it can be problematic during the pre-symptomatic or early asymptomatic phase of infection.^10^ As demonstrated by our longitudinal studies (**Figure 2D**), an individual can be positive by PCR but negative by antigen test for one or two days before testing positive. A negative result delivered at this stage in the course of infection could offer false security to someone who is about to become highly infectious. Furthermore, with the time window of positive results narrower than for PCR testing, relatively small differences in test sensitivities can translate to capturing or missing potentially infectious cases. We therefore recommend that regular testing be emphasised, and that tests are deployed in populations where the limitations of these tests are understood and/or manageable.

In certain in-patient situations, LFDs can also be used to make early, rapid decisions about patient management, with appropriate isolation pending confirmatory SARS-CoV-2 PCR testing. This approach has recently been successful in hospital LFD pilot studies, with the use of such devices preventing the cohorting of asymptomatic and/or infectious individuals with uninfected patients while awaiting PCR results.^26^ At the other end of the disease course, LFDs could also be useful for determining if persistently PCR positive individuals pose a transmission risk, potentially in tandem with rapid antibody testing.^27^

Although the LFDs are very easy to use, the correct sampling, reading and interpretation of the result are essential to their success in mass screening situations.^1^ In particular, one must take into account training and familiarity with swabbing when deploying devices to the general public as compared with a trained healthcare worker in a hospital/clinic-based setting. It is also easy to underestimate the importance of correctly recognising a positive band. We found that some tests gave clearer results than other, as illustrated by the number of borderline results seen in Figure 1 and 2. Removing this element of subjectivity, for example through the use of an smartphone application to read/capture the LFD result, could improve success rates.

Our data support the judicious use of rapid antigen LFDs – not to replace PCR testing, but to supplement the current testing capacity and rapidly identify infected individuals in situations where they would otherwise go undetected. Although sensitivity is lower than PCR-based testing, the rapid turnaround of these tests, their versatility in terms of cost and portability, and their utility in disrupting transmission chains originating from infectious asymptomatic individuals, outweighs the risk of missing positive cases.

## Data Availability

All relevant data are within the manuscript.

## Acknowledgments

We thank the following sources for donation of test kits: the manufacturers of Spring, SureScreen Fluorescent and SureScreen Visual, Encode, Innova and E25Bio.

We are extremely grateful to all patients and staff at St Thomas’ Hospital who participated in this study.

## Contributors

SP, RB, LS, BM, GN, SD, MHM, JDE, SJDN, RPG conceived the study and study design. SP, RB, BM, LS, MTKI, TX, AM, RPG collated the data. LS, BM, TX, AM, GN, SD supervised specimen collection. MJL and KJD generated reagents. SP and RPG analysed the data. SP, SJDN, and RPG wrote the first draft of the manuscript. All authors critically reviewed the manuscript. The corresponding author attests that all listed authors meet authorship criteria and that no others meeting the criteria have been omitted.

## Data Sharing

All relevant data are within the manuscript (and its supporting information files).

